# Genetic architecture of the personality meta-traits – stability and plasticity – and their overlap with psychopathology

**DOI:** 10.64898/2026.03.18.26348729

**Authors:** Liam J. Veltman, S. Hong Lee, Beben Benyamin, 23andMe Research Team, Sarah Cohen-Woods, Elina Hyppönen, David Stacey

## Abstract

The Five-Factor model (FFM) personality traits (agreeableness, conscientiousness, extraversion, neuroticism and openness) capture stable individual differences in thinking, feeling and behaviour. It has been shown that the FFM traits share variance through two-higher order “meta-traits”, stability and plasticity. It remains unknown, however, whether these meta-traits can capture the shared genetic architecture of the FFM personality traits. Here we combined recent genome-wide association study (GWAS) summary statistics with data from the 23andMe Research Institute (European ancestry; total N = 279,240-682,707) and applied Genomic Structural Equation Modelling, identifying two latent genetic factors consistent with stability and plasticity. We then performed a multivariate GWAS on these factors to capture genetic loci with shared effects across the FFM traits, identifying 81 and 13 independent genome-wide significant loci for stability and plasticity respectively. Transcriptome-wide and cell-type enrichment analyses prioritised candidate effector genes and indicated broad brain involvement for personality traits. Finally, genetic correlation and pleiotropy-aware Mendelian randomisation analyses provided genetic evidence suggesting bi-directional links between stability and psychopathology: higher genetic propensity for stability was protective against psychopathology, whereas greater genetic liability to psychopathology was associated with reduced stability. These results enhance our understanding of the shared genetic architecture underlying personality traits and their overlap with psychopathology.

## Introduction

Personality is a fundamental characteristic of humans, reflecting the enduring patterns of thinking, feeling and behaving which shape how individuals interact with the world around them. The broad individual differences in human personality can be captured by the Five Factor Model (FFM) or ‘Big 5’ personality traits, which consist of agreeableness (vs. antagonism), measuring compassion and cooperativity; conscientiousness (vs. undependability), capturing order and discipline; extraversion (vs. introversion) capturing talkativeness and assertiveness; neuroticism (vs. emotional stability) capturing sensitivity to negative emotion; and openness (vs. closedness), measuring intellectual curiosity and creativity^1^. As personality traits have been associated with many important life outcomes such as career choice^2^, educational attainment^3^, disease burden^4^ and longevity^5^, and show overlap with psychopathology^6^, insight into the genetic and biological underpinnings of personality will be essential to further our understanding of human behaviour.

Cybernetic Big Five Theory (CB5T) posits that the individual differences in human personality captured by the FFM reflect variation in the parameters of goal-directed, cybernetic mechanisms in the brain^7,8^. At a higher-order level, these domains cohere into two “meta-traits”: stability and plasticity, which capture the shared variance across the FFM traits^9,10^. Stability (agreeableness, conscientiousness and low neuroticism) captures individuals’ capacity to resist perturbation and maintain order across social, emotional and motivational systems^10^. Plasticity (extraversion and openness) on the other hand indexes one’s capacity to explore, learn and flexibly engage with novelty across behaviour and cognition^10^. Although CB5T offers a promising bridge to broader predictive coding frameworks in computational neuroscience^8^, direct biological evidence remains limited and indirect,^11^ due in large part to a lack of scalable research tools capable of teasing apart complex and nuanced brain mechanisms in situ. Recent advances in large-scale genomic datasets and multivariate statistical tools creates unprecedented opportunity to test whether these meta-traits reflect genetically coherent dimensions and to map the biological pathways that underpin human personality.

Links between personality traits and psychopathology have long been noted, with dimensional nosologies such as HiTOP noting the fuzzy boundaries between disposition and disorder^6,12,13^. The FFM traits which comprise stability are broadly protective of psychopathology; high agreeableness, high conscientiousness and low neuroticism are negatively associated with psychopathology with this effect being most pronounced for low neuroticism^14,15^. Extraversion shows facet specific patterns, with communal extraversion typically related with lower levels of internalising symptoms, whereas agentic extraversion tends to associate positively with externalising behaviours^16^. Finally, aspects of openness are positively linked with psychosis-spectrum traits^17^ and also share genetic loci with schizophrenia^18^. Although the genetic overlap between the FFM traits and psychiatric conditions has been characterised previously^19–21^, it remains unknown whether these genetic correlations can be captured by shared dimensions of genetic overlap between personality traits such as stability and plasticity.

Apart from neuroticism, genome-wide association studies (GWAS) of the FFM personality traits were historically underpowered relative to other psychiatric traits. Recently, Gupta et al.^20^ substantially boosted the sample size for all five FFM traits in a cross-ancestry GWAS meta-analysis and reported 208, 14, 3, 2 and 7 genome-wide significant loci associated with neuroticism, extraversion, agreeableness, conscientiousness and openness respectively, expanding upon previous efforts^19,21–24^. Beyond increased power, multivariate tools such as Genomic Structural Equation Modelling (SEM) and functional extensions (e.g. transcriptome-wide SEM and stratified Genomic SEM) enable modelling of latent genetic dimensions and prioritisation of biological pathways underlying shared risk^25–27^. These multivariate approaches have successfully identified shared dimensions of genetic risk and convergent biology across psychiatric conditions^26,28^. Here we apply these same methods to the recently updated personality GWAS to investigate whether the FFM meta-traits stability and plasticity are reflected in the genome and to highlight shared biology across the FFM personality traits.

In this work, we conduct a multivariate genomic investigation into the FFM meta-traits of stability and plasticity. We firstly combined the European GWAS summary statistics from the recent Gupta et al.^20^ work with summary statistics from the 23andMe Research Institute^19^ and conducted a GWAS meta-analysis across ∼280,000-680,000 individuals for each of the FFM personality traits. Next, using Genomic SEM, we identified 2 latent genetic factors consistent with the meta-traits stability and plasticity and performed a multivariate GWAS of these latent factors, identifying genetic loci having shared effects across the FFM traits. We then conducted transcriptome-wide association studies (TWAS) and cell type enrichment analyses to highlight potential biological pathways underpinning the FFM traits/meta-traits. Finally, we quantified genetic overlap between the FFM traits/meta-traits and 11 psychiatric conditions using genetic correlations and tested for directional associations using bi-directional, pleiotropy aware Mendelian randomisation (MR) analyses.

## Results

### GWAS meta-analysis of FFM personality traits

To increase power for downstream analyses, we first meta-analysed all currently available European GWASs for the FFM personality traits by combining the 23andMe Research Institute summary statistics from Lo et al.^19^ with those from Gupta et al.^20^ using METAL^29^. Results of our meta-analyses are summarised below in Table 1.

**Table 1.**
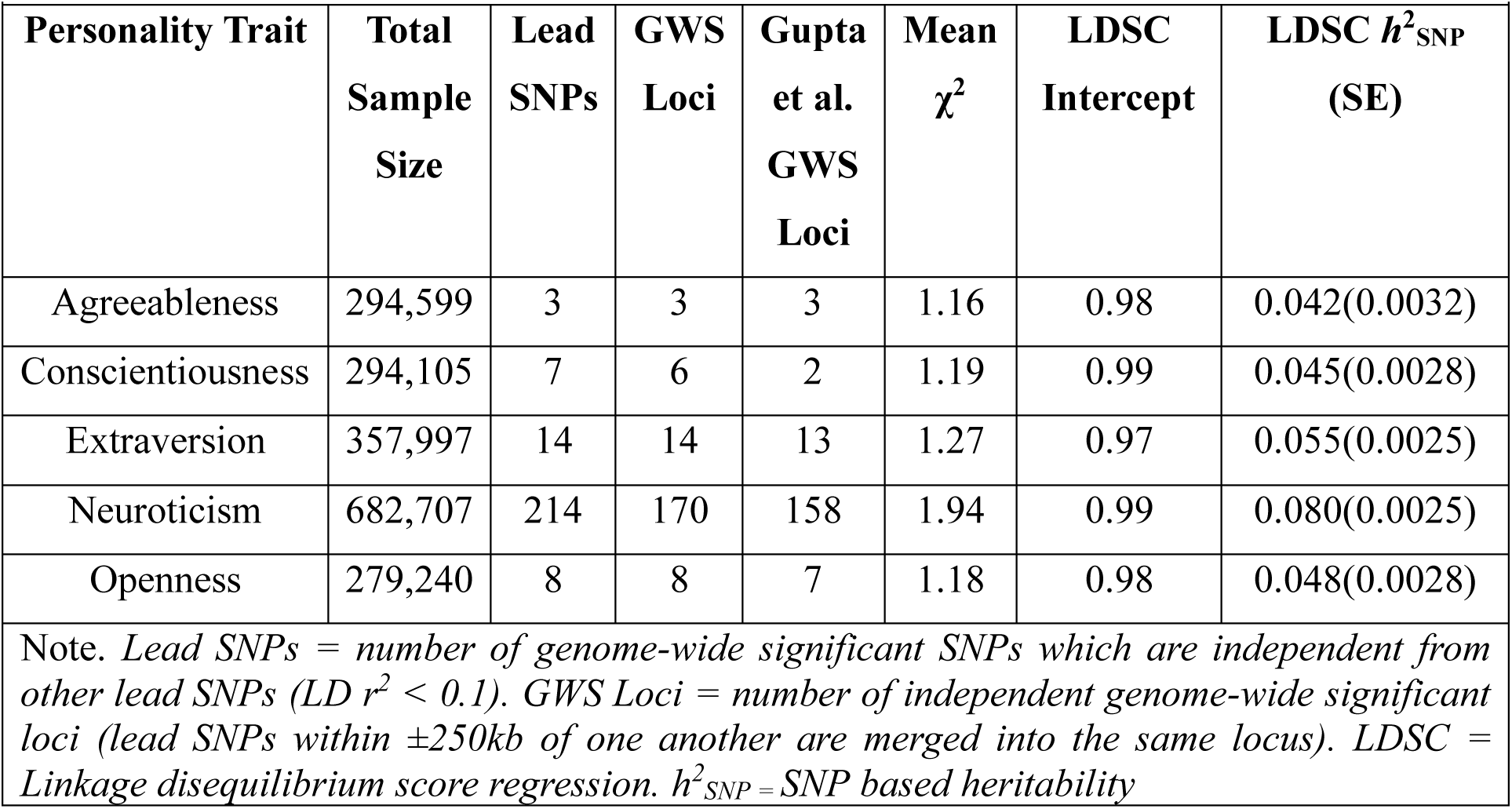
Summary of Results for Genome-Wide Association Study Meta-Analyses of the Five Factor Model Personality Traits.

| Personality Trait | Total Sample Size | Lead SNPs | GWS Loci | Gupta et al. GWS Loci | Mean $\chi^2$ | LDSC Intercept | LDSC $h^2_{\text{SNP}}$ (SE) |
| --- | --- | --- | --- | --- | --- | --- | --- |
| Agreeableness | 294,599 | 3 | 3 | 3 | 1.16 | 0.98 | 0.042(0.0032) |
| Conscientiousness | 294,105 | 7 | 6 | 2 | 1.19 | 0.99 | 0.045(0.0028) |
| Extraversion | 357,997 | 14 | 14 | 13 | 1.27 | 0.97 | 0.055(0.0025) |
| Neuroticism | 682,707 | 214 | 170 | 158 | 1.94 | 0.99 | 0.080(0.0025) |
| Openness | 279,240 | 8 | 8 | 7 | 1.18 | 0.98 | 0.048(0.0028) |
| Note. Lead SNPs = number of genome-wide significant SNPs which are independent from other lead SNPs ($LD r^2 < 0.1$). GWS Loci = number of independent genome-wide significant loci (lead SNPs within $\pm 250\text{kb}$ of one another are merged into the same locus). LDSC = Linkage disequilibrium score regression. $h^2_{\text{SNP}}$ = SNP based heritability | | | | | | | |

In total, 211 independent genome-wide significant loci (i.e. p < 5e^-8^), were identified in our univariate GWAS meta-analyses of the FFM personality traits. Apart from neuroticism, for which 2 loci from the Gupta et al. meta-analysis were no longer genome-wide significant, all loci identified in the previous meta-analysis were replicated. Our meta-analyses also identified 4, 1, 14 and 1 novel loci which were not identified in Gupta et al. for conscientiousness, extraversion, neuroticism and openness respectively. Summary of genome-wide significant loci and mapped genes for each FFM trait can be found in Supplementary Tables 1-5. Summary statistics for the top 10,000 SNPs for each FFM trait can be found in Supplementary Tables 6-10. Manhattan and q-q plots for each GWAS can be found in Supplementary Note 1 and 2.

### Genetic Architecture of the FFM Meta-Traits

To investigate whether there is a genetic basis for the FFM meta-traits stability and plasticity^9,10^, we first used LDSC regression^30^ to estimate genetic correlations between the FFM personality traits (Figure 1 A). For ease of interpretation and consistency with prior literature, the effect estimate column for neuroticism was multiplied by −1 to produce inverted neuroticism which is used throughout subsequent analyses^10^. The largest genetic correlations observed were between extraversion and openness (0.39) followed by agreeableness and inverted neuroticism (0.33).

**Figure 1.**
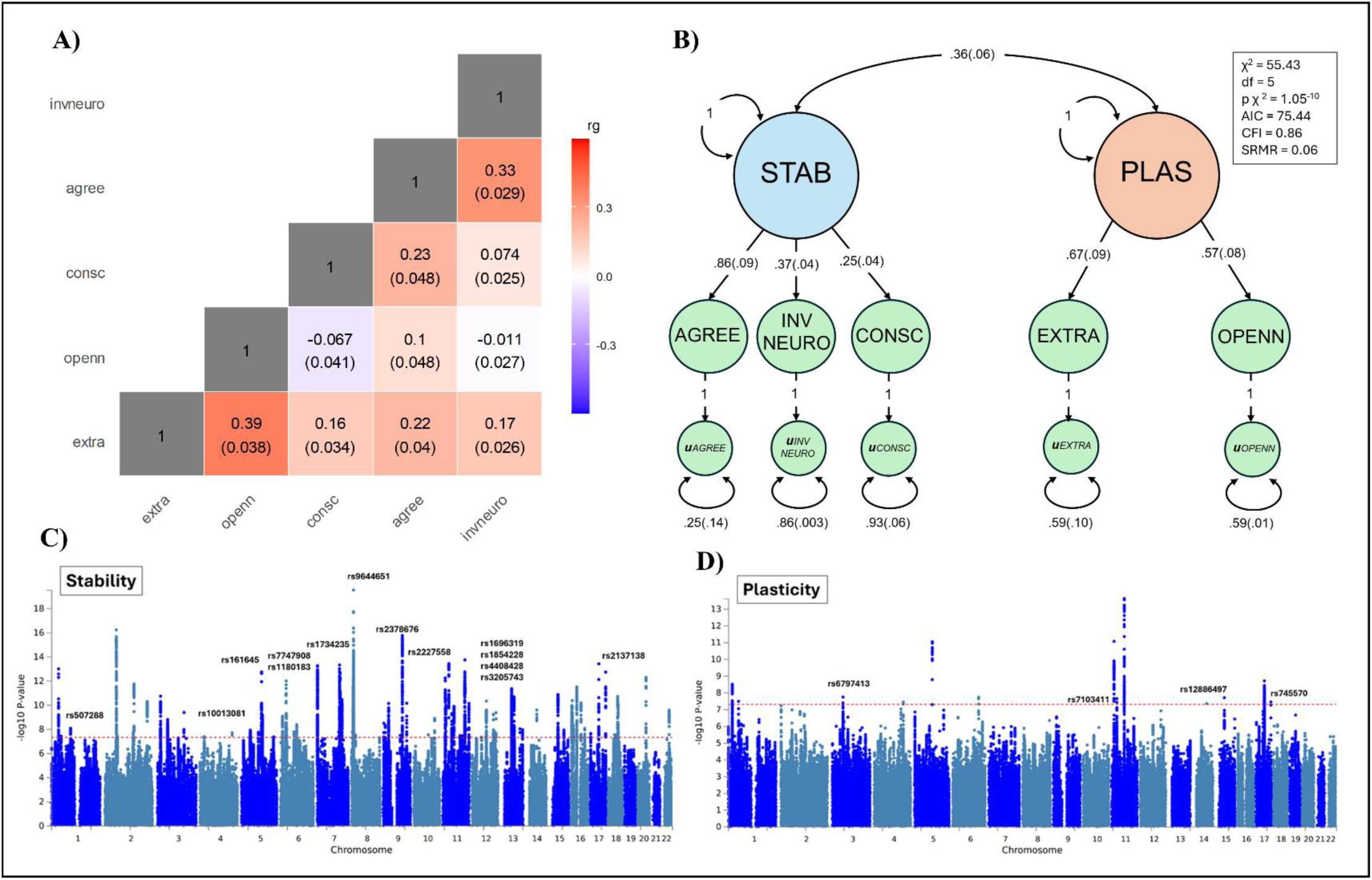
A) Heatmap of genetic correlation (rg) estimates between the Five Factor Model personality traits. B) Factor loadings and model fit for the meta-trait model of the Five Factor Model personality traits. C) & D) Manhattan plots of genome-wide association study statistics for stability (C) and plasticity (D). Dashed lines correspond to the genome-wide significance threshold (p = 5e^-8^). Labels correspond to the lead SNP at each genome-wide significant locus that was not identified in the univariate GWAS meta-analyses. For readability, labels are placed above the chromosome on which each locus is located. Note. agree = agreeableness, invneuro = inverted neuroticism, consc = conscientiousness, extra = extraversion, openn = openness, stab = stability, plas = plasticity, standard errors in parentheses.

We next conducted an exploratory factor analysis (EFA) on the genetic covariance matrix of the FFM traits. The EFA supported a 2-factor solution which was consistent with the FFM meta-traits. The first factor, interpreted as stability, was characterised by loadings from agreeableness (0.64), inverted neuroticism (0.46) and conscientiousness (0.35). The second factor, interpreted as plasticity, was characterised by loadings from openness (0.99) and extraversion (0.43). This pattern of loadings for stability was weaker than what is typically observed at the phenotypic level, however the general pattern was still observed^9^. Full results from the EFA are presented in Supplementary Table 11.

This two-factor solution was carried forward for confirmatory factor analysis (CFA) using Genomic SEM^25^. We also tested a one-factor model to investigate evidence for a general genetic factor of personality^31^; however, this model indicated poor fit to the data (AIC = 144.89, CFI = 0.67, SRMR = 0.093). The initial two-factor CFA indicated good fit (AIC = 51.7, CFI = 0.92, SRMR = 0.037). However, parameter estimates for extraversion were out of bounds (Heywood case^32^), reflecting the fact that the plasticity factor was defined by only two indicators. Latent factors with only two indicators are not fully identified and therefore require additional constraints. To address this issue, we tested a series of alternative constrained CFA models (Supplementary Table 12). The best-fitting model constrained the residual variances of extraversion and openness to equality, which successfully identified the plasticity factor (Figure 1 B). This model displayed adequate fit (AIC = 75.44, CFI = 0.86, SRMR = 0.056) and was retained for all subsequent analyses. These results support a genetic basis for the personality meta-traits stability and plasticity, which reflect the shared genetic architecture among the FFM personality traits.

We next conducted a GWAS of the FFM meta-traits to capture genetic loci having shared effects across the FFM personality traits by estimating SNP effects on the latent factors of stability and plasticity in Genomic SEM^25^. For stability, we identified 81 independent genome-wide significant loci, 14 of which were not identified in our univariate FFM trait GWASs (Figure 1 C). Notably, one novel locus for stability was mapped to *NTRK2* (chromosome 9, position = 87585184-87664625, rs2378676) which encodes the BDNF receptor TrkB, implicated in antidepressant action and synaptic plasticity^33^. For plasticity, we identified 13 independent genome-wide significant loci, 4 of which were not present in our original FFM trait GWASs (Figure 1 D). Interestingly, one of the novel loci for plasticity (chromosome 11, position = 27541623-27742447, rs7103411) was mapped to *BDNF*, a key regulator of synaptic plasticity implicated in learning, memory and many psychiatric disorders^34^. A summary of all genome-wide significant loci for stability and plasticity can be found in Supplementary Tables 13 and 14. The top 10,000 SNPs for the meta-traits can be found in Supplementary Tables 15 and 16.

Genomic SEM also computes a SNP level heterogeneity metric (QSNP) which identifies SNPs which have patterns of effects better explained by an independent pathways model rather than a latent factor. For stability, we identified 51 lead QSNPs, and for plasticity we identified 7. Lead QSNPs and their LD partners are presented in Supplementary Tables 17 and 18.

### Cell Type Enrichment

To investigate whether the genetic signal from our FFM personality trait GWASs were enriched in specific brain cell types, we used stratified LD score regression (S-LDSC) and estimated SNP-heritability enrichment across the 31 brain cell type annotations from Yao et al.^35^. These annotations are for the top decile of genes expressed for each of the 31 brain cell superclusters identified in Siletti et al.^36^. We also estimated heritability enrichment across the baselineLD v2.2 annotations which capture a spectrum of genomic features such as regulatory elements and conservation metrics^37^. For the FFM meta-traits, we used stratified Genomic SEM^26^ to investigate SNP heritability enrichment across these same annotations for stability and plasticity. A Benjamini-Hochberg correction for multiple testing was applied to all results from our enrichment analyses. Full results from the S-LDSC and stratified Genomic SEM analyses are presented in Supplementary Tables 19-25.

Overall, the FFM personality traits primarily show enrichment across both cortical and sub-cortical neurones, whilst glial and support cells do not show enrichment despite a few exceptions (Figure 2). Agreeableness showed enrichment in fewer cell types than the other FFM traits, likely due to the agreeableness GWAS having fewer genome-wide significant loci. Despite this, the broader pattern of enrichment for the FFM traits suggests a brain wide genetic contribution to personality and implicates cortical and sub-cortical neurones as being principally relevant for personality traits.

**Figure 2.**
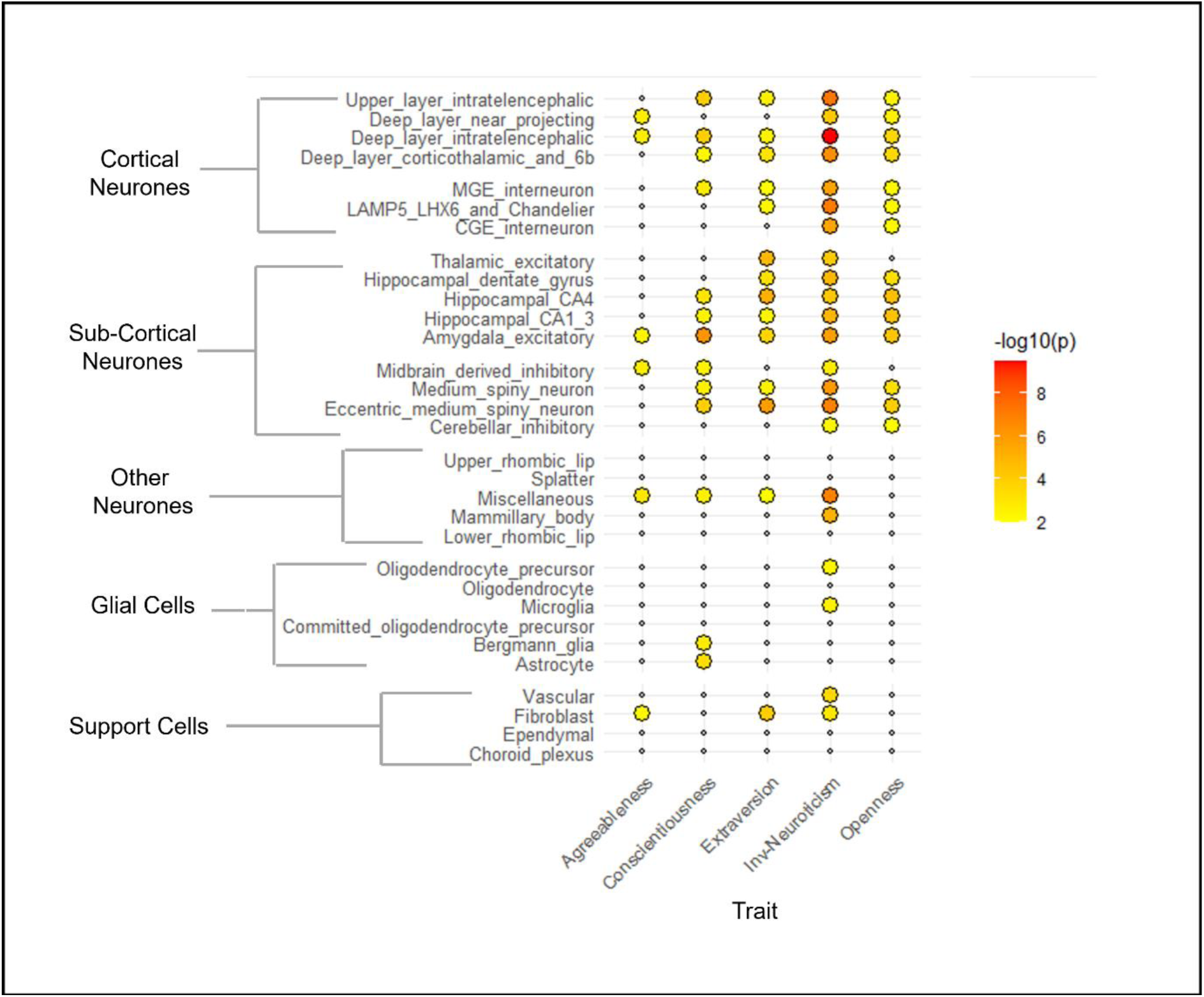
Bubble plot heatmap showing -log10 enrichment p-values across 31 brain cell types for each of the Five Factor Model personality traits. Note. The groupings of the superclusters from Siletti et al (cortical, subcortical and others) were used to assist interpretation. Coloured dots represent enrichments which had FDR_BH_ < 0.05. None of the cell type annotations had FDR_BH_ < 0.05 for stability or plasticity and hence were omitted from the plot.

For both meta-traits, no brain cell annotations displayed significant enrichment in the stratified Genomic SEM analyses after FDR correction. Several of the baseline annotations also produced warnings about negative residual variances, suggesting that our FFM trait GWASs are currently underpowered for stratified Genomic SEM. For stability, the only significantly enriched annotation was for highly conserved regions of the genome (Genomic Evolutionary Rate Profiling [*GERP.RSsup4*], FDR_BH_ = 0.02), which are thought to be under strong negative selection and integral to the overall functioning of an organism. Interestingly, conserved regions have also been shown to be enriched for general psychopathology^26^ suggesting a potential genetic pathway linking trait stability and general psychopathology risk. Although no annotations were significant for plasticity after FDR correction, the two strongest signals were for amygdala excitatory neurones and eccentric medium spiny neurones (p = 0.004 & 0.007 respectively; FDR_BH_ = 0.14), both of which are implicated in novelty detection and salience attribution^38,39^ which are key components of trait plasticity^7^.

### Transcriptome Wide Association Study

To identify potential effector gene candidates, we used the summary statistics generated from our FFM GWAS to conduct a TWAS in the 13 available brain tissues and whole blood from GTEx v8^40^ using FUSION^41^. We then used these fusion results to conduct a TWAS of the FFM meta-traits using T-SEM^27^. A strict Bonferroni correction for multiple testing was applied (p < 0.05/number of gene-tissue models) in addition to a more permissive Benjamini-Hochberg correction (FDR_BH_ < 0.05) to identify suggestive gene-trait associations. T-SEM also computes a Q-trait metric which, like the QSNP metric, identifies gene-trait associations with patterns of effects better explained by an independent pathways model rather than a latent factor. Results for the meta-traits were filtered to exclude gene-trait associations with a Bonferroni corrected Q-trait p-value < 0.05 prior to interpretation. A summary of gene-trait associations is presented in Table 2. Suggestive gene-trait associations broken down by tissue are presented in Figure 3 A, and a summary of all TWAS hits is presented in Supplementary Tables 26-32. Full TWAS results are presented in Supplementary Tables 33-39.

**Figure 3.**
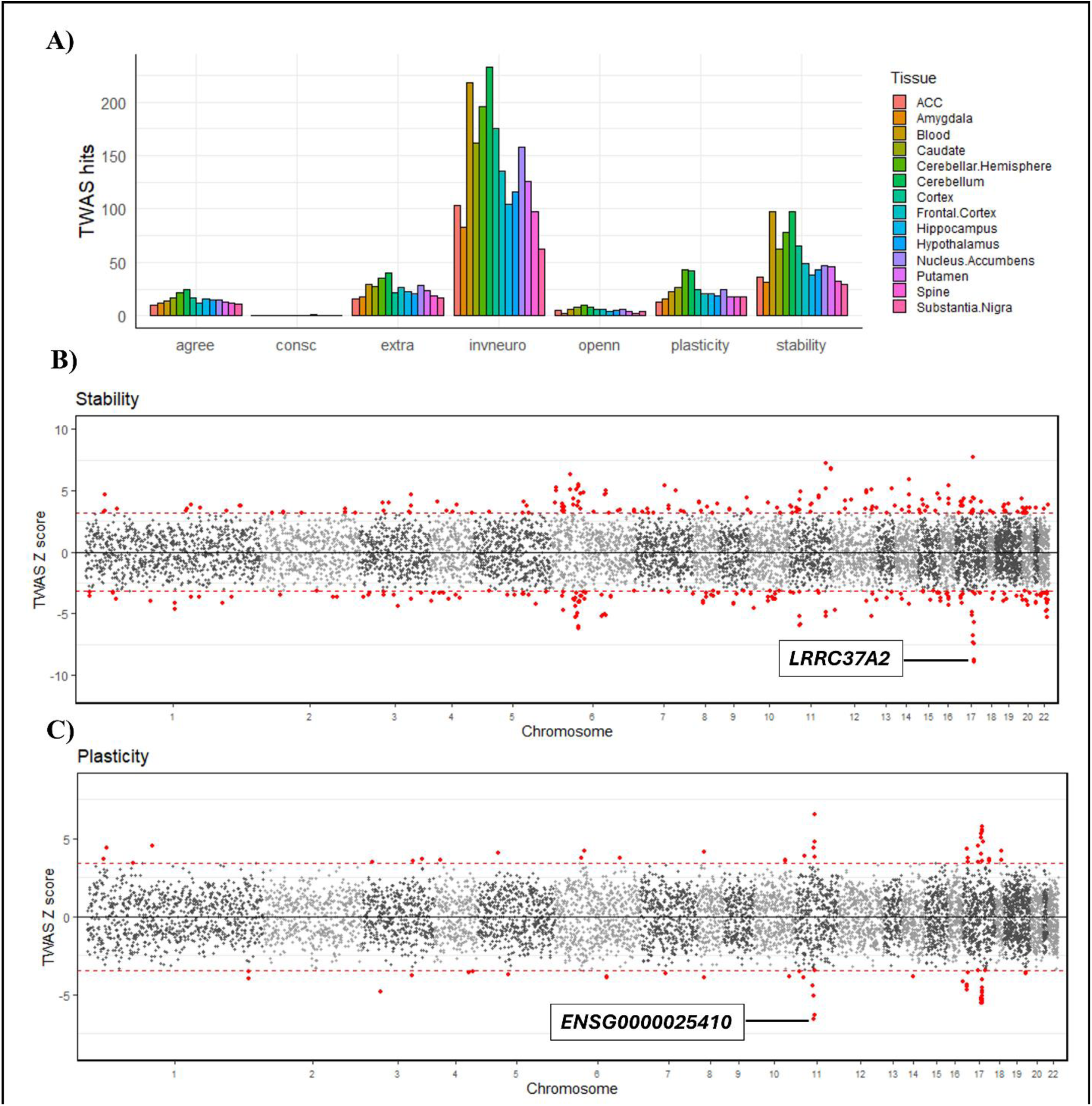
A) Bar chart of the number of suggestive gene-trait associations from our transcriptome wide analyses in each of the GTEx v8 sub-tissues tested for the Five Factor Model personality traits/meta-traits. B) & C) Scatterplots of gene-trait association Z-scores for the meta-traits stability and plasticity. Labels correspond to the strongest gene-trait association observed. Dashed lines correspond to the suggestive FDR_BH_ < 0.05 threshold for each meta-trait. Note. agree = agreeableness, consc = conscientiousness, extra = extraversion, invneuro = inverted neuroticism, openn = openness, ACC = anterior cingulate cortex.

**Table 2.** Summary of Significant and Suggestive Gene-Trait Associations across Transcriptome Wide Association Analyses for the Five Factor Model Personality Traits and Meta-Traits.

|  |  |  |
| --- | --- | --- |
| Personality Trait | Number of gene-models |  |
| | $\text{FDR}_{\text{BH}} < 0.05$ (suggestive) | Bonferroni corrected $p < 0.05$ |
| Agreeableness | 45 | 0 |
| Conscientiousness | 1 | 1 |
| Extraversion | 85 | 24 |
| Inverted Neuroticism | 782 | 159 |
| Openness | 34 | 9 |
| Stability | 363 | 51 |
| Plasticity | 88 | 28 |
| *Note. Counts correspond to gene-trait associations which were significant/suggestive in at least 1 of the 13 sub-tissues tested.* |  |  |

With the exception of conscientiousness, for which the only significant gene-trait association was *PLCH1* in the hypothalamus, significant/suggestive gene-trait associations were present throughout all brain tissues tested for the FFM traits/meta-traits (Figure 3 A). Notably, the strongest signal for plasticity was *ENSG00000254510* in the nucleus accumbens (Figure 3 B), a long non-coding RNA located ∼ 4000bp upstream of *NPAS4*, an activity dependent transcription factor involved in synaptic plasticity^42^. The nucleus accumbens is also part of the mesolimbic dopamine pathway which has been theorised to contribute to the meta-trait plasticity^7^. The strongest signal for trait stability was *LRRC37A2* (Figure 3 C), which has been suggested to play a role in foetal brain development^43^ and has previously been implicated in other cognitive traits and neurological conditions^44,45^.

### Genetic Correlations Between FFM Personality Traits and Psychiatric Conditions

We then sought to investigate the genetic overlap between the FFM personality traits and 11 different psychiatric conditions^46–56^ using LDSC regression^30^ (a summary of the psychiatric condition GWAS used in the genetic correlation analyses are presented in Supplementary Note 3; full results from genetic correlation analyses are presented in Supplementary Tables 40 and 41). Genetic correlations for the FFM traits and meta-traits are visualised in Figure 4 A and B respectively.

**Figure 4.**
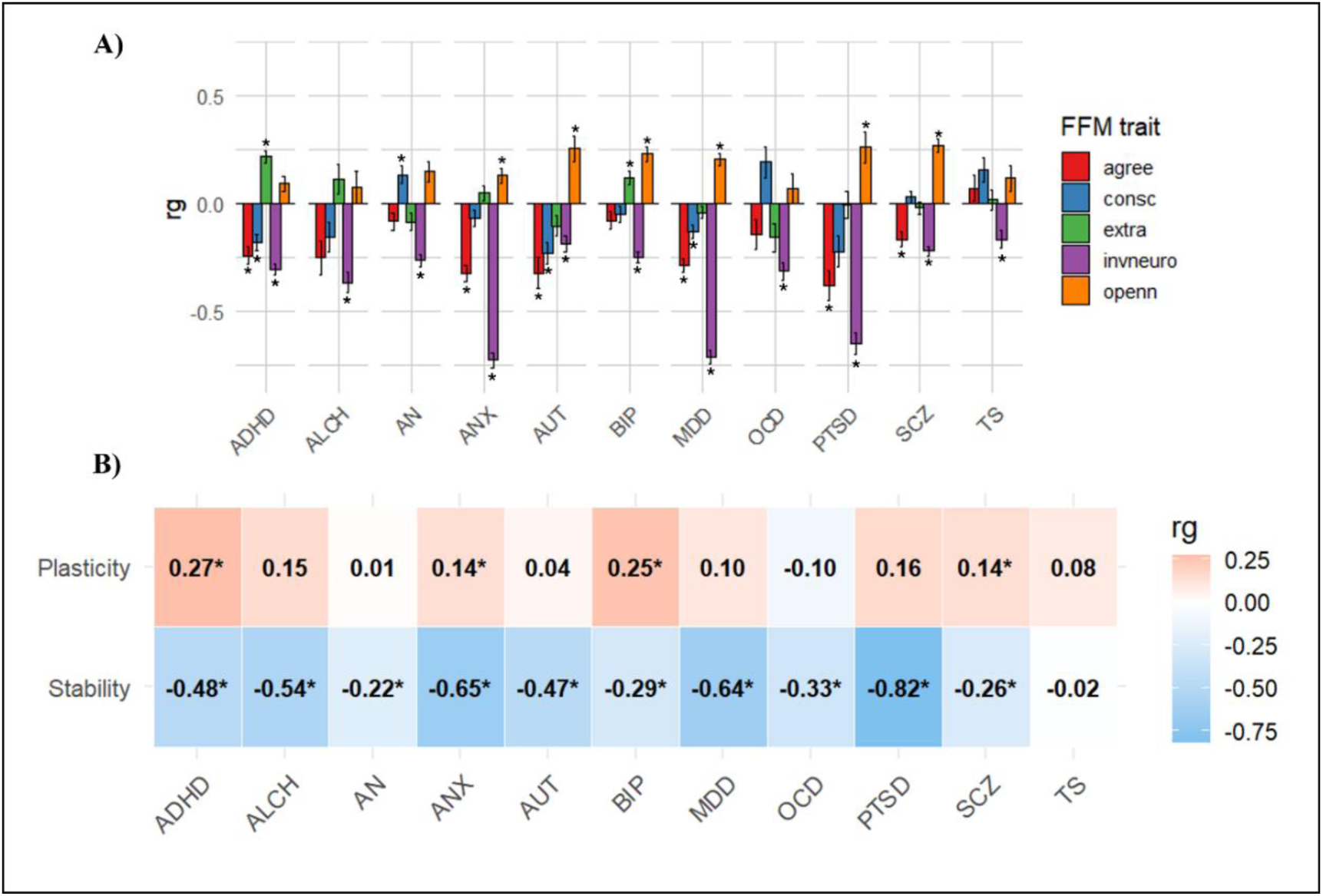
A) Bar chart of genetic correlations (rg) between the FFM personality traits and 11 psychiatric disorders. Note. * = significant at Bonferroni correction for 55 tests (p < 0.0009), error bars represent standard errors. B) Heatmap of genetic correlations between the FFM meta-traits and 11 psychiatric conditions. Note. * = significant at Bonferroni correction for 22 tests (p < 0.0023). ADHD = attention deficit hyperactivity disorder, ALCH = problematic alcohol use, AN= anorexia nervosa, ANX = anxiety, AUT = autism spectrum disorder, BIP = bi-polar disorder, MDD = major depressive disorder, OCD = obsessive compulsive disorder, PTSD = post-traumatic stress disorder, SCZ = schizophrenia, TS = Tourette syndrome, agree = agreeableness, consc = conscientiousness, extra = extraversion, invneuro = inverted neuroticism, openn = openness.

As has been demonstrated previously, inverted neuroticism was consistently negatively associated with all psychiatric conditions, with the strongest correlation being seen for ANX, MDD and PTSD (rg = −0.73, −0.71 & −0.65 respectively). Agreeableness showed moderate negative associations with ADHD, ANX, AUT, MDD, PTSD and SCZ (rg ranging from −0.17 to −0.38). Conscientiousness had a small positive association with AN and small negative associations with ADHD, AUT and MDD (rg = −0.18, −0.23 & −0.13 respectively). Extraversion had a moderate positive association with ADHD (rg = 0.22) and small positive association with BIP (rg = 0.12). Finally, openness displayed a small positive association with ANX (rg = 0.13) and moderate positive associations with AUT, BIP, MDD, PTSD and SCZ (rg ranging from 0.20 to 0.27). Interestingly, all three traits which define stability (agreeableness, conscientiousness and inverted neuroticism) largely show negative associations with the psychiatric conditions tested, suggesting that stability may be able to capture these shared associations.

We next used Genomic SEM^25^ to estimate the genetic overlap between the FFM meta-traits, which capture the shared genetic architecture of the FFM personality traits, and these same psychiatric conditions (Figure 4 B). As was suggested by the pattern of effects for the FFM traits, stability showed consistent negative associations with all psychiatric conditions besides TS. This pattern was most pronounced for the internalising conditions (ANX, MDD, PTSD). Plasticity on the other hand displayed moderate positive associations with both ADHD and BIP and small positive associations with ANX and SCZ but otherwise did not display genetic overlap with other psychiatric conditions.

### MR analyses

To investigate genetic evidence for a causal relationship between FFM personality traits and psychiatric conditions, we next conducted bi-directional MR analyses for all personality traits and psychiatric conditions which had significant genetic overlap in our genetic correlation analyses. We used CAUSE^57^ as the method specifically compares a causal model to a shared pleiotropy model, which is essential in the context of personality traits and psychiatric conditions where the boundaries between these traits are unclear^58^. We considered nominally significant (CAUSE p < 0.05) results as suggestive of causality and Benjamini-Hochberg corrected p-values < 0.05 as evidence of a causal relationship. All results with CAUSE p < 0.05 are presented in Table 3, and full results from all CAUSE analyses are presented in Supplementary Tables 42-50. Forest plots summarising reverse causal effect estimates for psychiatric conditions on personality traits are presented in Supplementary Note 4.

**Table 3.**
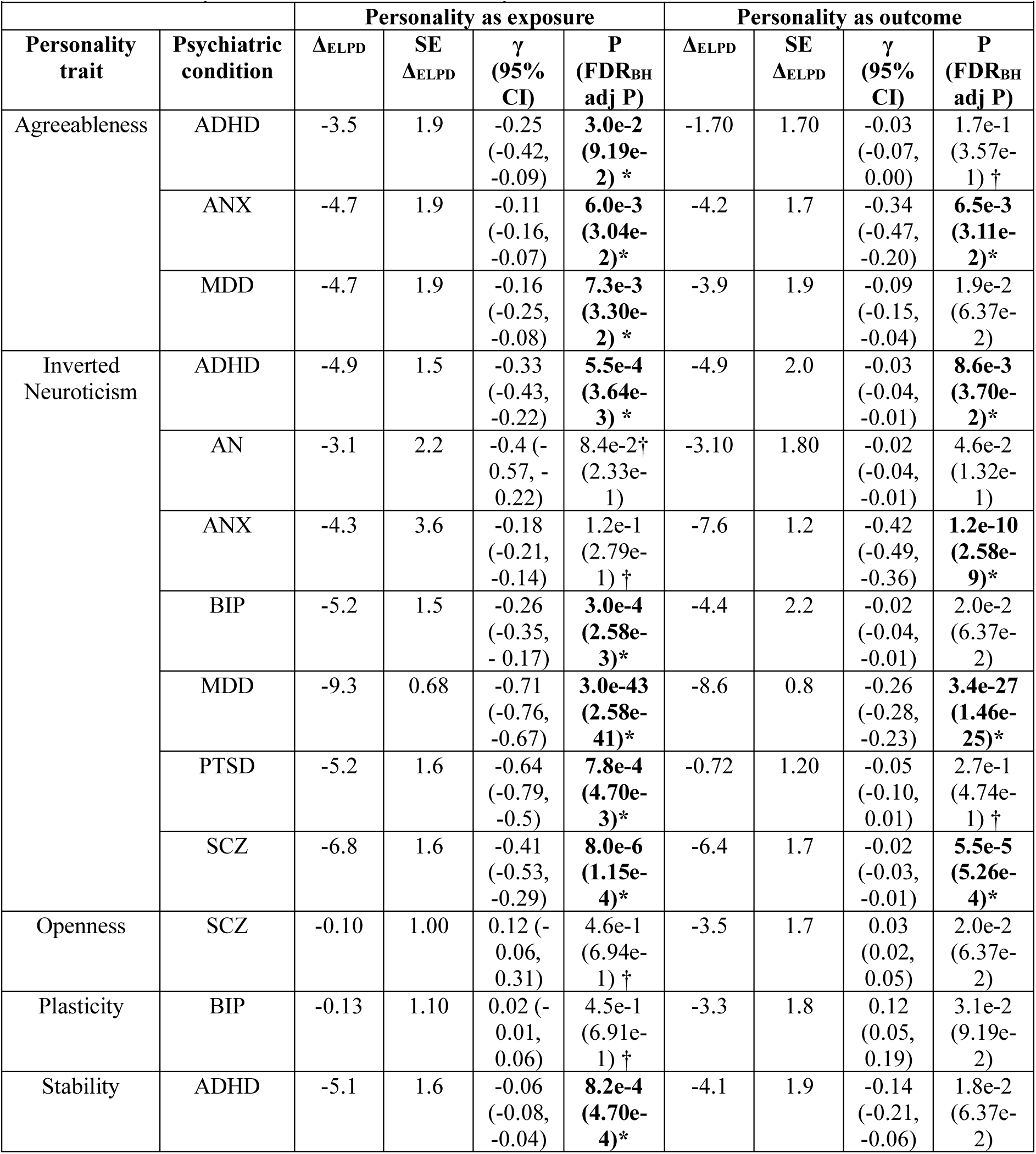
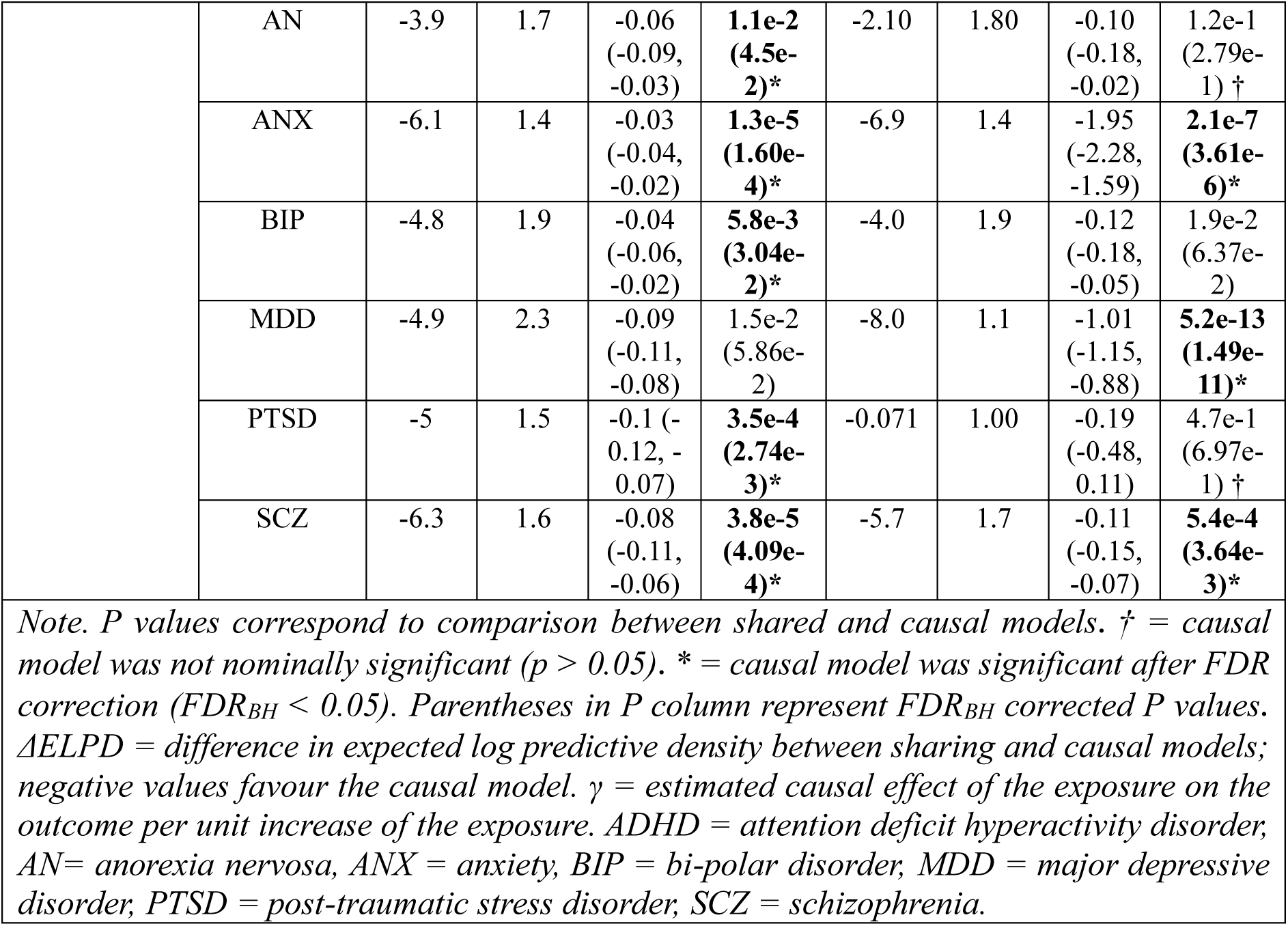
Summary of CAUSE Mendelian Randomisation Results between the Five factor Model Personality Traits/Meta-Traits and Psychiatric Conditions.

Agreeableness, inverted neuroticism and the meta-trait stability all displayed genetic evidence for a negative effect on psychiatric conditions (visualised in Figure 5). Genetic propensity for high agreeableness was protective of ANX and MDD in addition to there being suggestive evidence for a protective effect on ADHD. There was also suggestive evidence of a reverse causal effect for ANX and MDD on agreeableness, with genetic risk for these psychiatric conditions being negatively associated with agreeableness.

**Figure 5.**
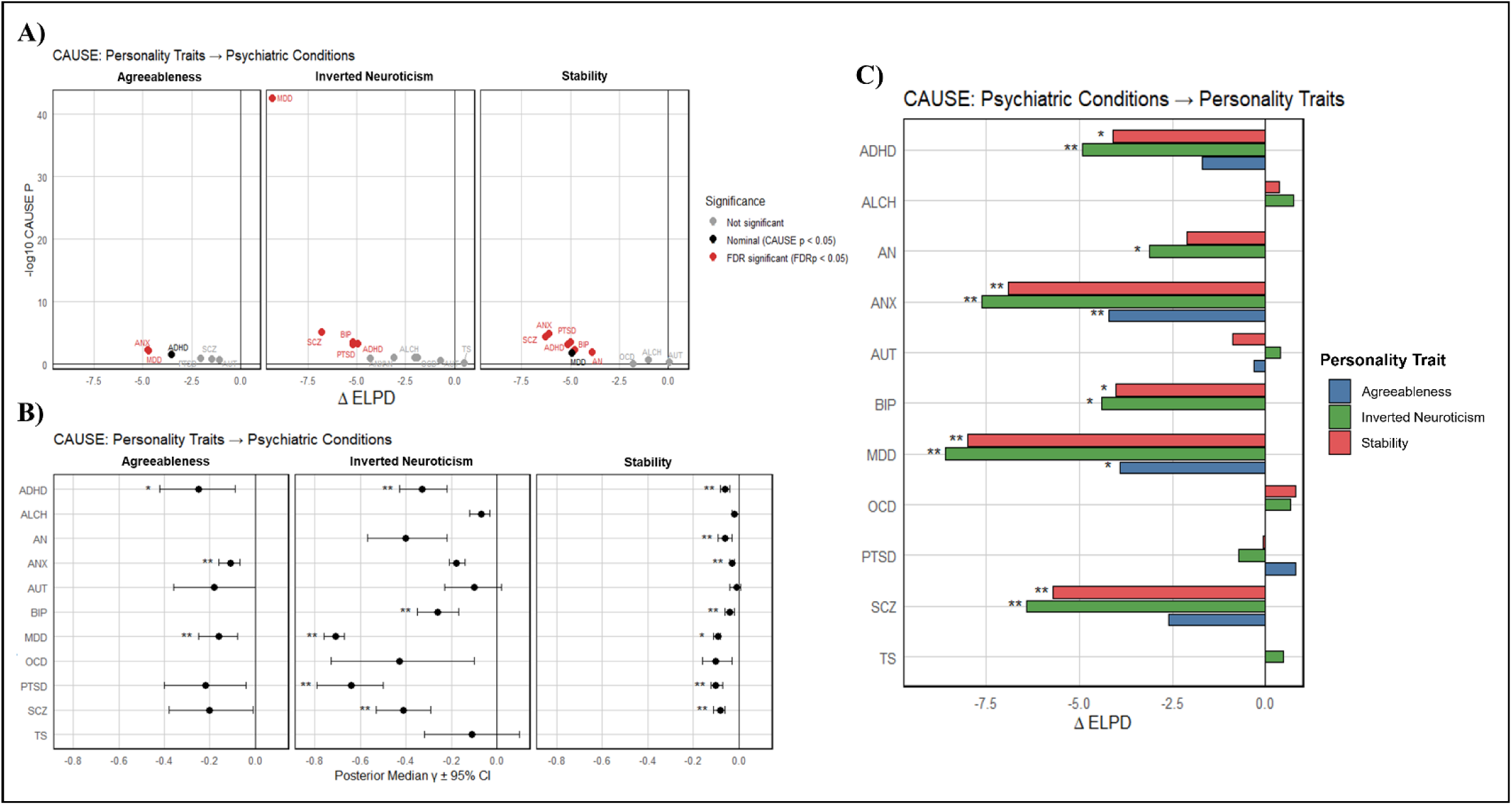
A) Scatter plots summarising CAUSE models for agreeableness, inverted neuroticism and stability as exposures and psychiatric conditions as outcomes. Each point represents a personality-psychiatric condition pair. The x axis corresponds to the change in expected log pointwise posterior density (ΔELPD) between causal and shared pleiotropy models, with negative values indicating a causal model fit better than a sharing model and vice versa. - log10(CAUSE p) values are plotted on the y axis. Black dots represent nominally significant results (CAUSE p < 0.05), and red dots represent results with FDR_BH_ < 0.05. B) Forest plots showing causal effect estimates of agreeableness, inverted neuroticism and stability on psychiatric conditions (posterior median γ ± 95% CI). C) Bar charts summarising CAUSE models for psychiatric traits as exposure and agreeableness, inverted neuroticism and stability as outcomes. Again, the x axis corresponds to the ΔELPD between causal and shared pleiotropy models. Note: Effect sizes are scaled to the exposure trait and indicate relative causal influence, not absolute magnitude. * = CAUSE p < 0.05, ** FDR_BH_ < 0.05).

For inverted neuroticism, evidence of a negative effect was observed for ADHD, BIP, MDD, PTSD and SCZ. Interestingly, the causal model was not preferred over the shared model for ANX (CAUSE p = 0.12) suggesting shared pleiotropy accounts for much of the relationship between inverted neuroticism and ANX (Figure 5 A). In the reverse direction, ADHD, ANX, MDD and SCZ displayed a negative effect on inverted neuroticism (i.e. increased neuroticism), with there being suggestive evidence for AN and BIP as well (Figure 5 C; Supplementary Note 4).

Higher genetic liability for the meta-trait stability was negatively associated with multiple psychiatric conditions (ADHD, AN, ANX, BIP, PTSD, SCZ), however, there was only suggestive evidence of a protective effect of stability on MDD (CAUSE p = 0.015; FDR_BH_ = 0.059) (Figure 5 B). Again, in the reverse direction, ANX, MDD and SCZ displayed evidence of a negative effect on stability with suggestive effects also being observed for ADHD, AN and BIP (Figure 5 C; Supplementary Note 4).

Overall, both inverted neuroticism and the meta-trait stability show genetic evidence for protective effects across many psychiatric conditions, with a higher genetic propensity for these traits being negatively associated with psychopathology. There is also substantial evidence that psychiatric conditions exert negative effects on these personality traits, suggesting complex bi-directional pathways linking emotional stability and psychiatric conditions. Apart from suggestive positive effects of schizophrenia on openness and bipolar disorder on plasticity, there was no evidence for any other causal relationships for conscientiousness, extraversion, openness and plasticity, suggesting that genetic liability for these traits plays a limited role in psychiatric risk.

## Discussion

In the present study we integrated large-scale GWAS meta-analyses with multivariate modelling to test whether the FFM meta-traits of stability and plasticity reflect genetically coherent dimensions of personality. While the factor structure is weaker than what is observed in the phenotypic literature, our modelling indicated that the genetic covariance between the FFM traits was best described by two factors corresponding to stability (agreeableness, conscientiousness and inverted neuroticism) and plasticity (extraversion and openness), whereas a single general factor of personality model fit poorly. These results are consistent with CB5T^7^ which posits a biological basis to these higher order meta-traits which reflect individual differences in goal directed control and exploratory mediated belief updating. Moreover, our multivariate GWASs of the FFM meta-traits highlighted biologically plausible genetic loci beyond our univariate trait GWASs (e.g. *NTRK2* and *BDNF)*, further supporting the notion that the meta-traits reflect genetically coherent dimensions rather than measurement artefacts.

Within CB5T, the FFM meta-traits are conceptualised as high level ‘control parameters’ governing how individuals identify goals, select strategies for achieving them, and track progress towards desired states^7,8^. Stability broadly reflects the ability to pursue goals without disruption across emotional, social and motivational domains - captured by low neuroticism, agreeableness and conscientiousness respectively^7^. Conversely, plasticity captures individuals tendency to explore alternative goals and strategies both cognitively (indexed by openness), and behaviourally (indexed by extraversion)^7^. In this framework, genetic variants associated with stability likely contribute to differences in individuals’ broad ability to maintain coherent goal directed behaviour, whereas variants associated with plasticity contribute to general tendencies towards novelty seeking, exploration and goal updating.

Results from our TWAS/T-SEM analyses were consistent with diffuse genetic effects across both cortical and subcortical brain regions. There was at least suggestive evidence for gene-trait associations across each of the brain tissues tested for all FFM traits/meta-traits, except conscientiousness. One notable TWAS signal was for a lncRNA located ∼4kb upstream of *NPAS4* in the nucleus accumbens which emerged as our top signal for plasticity. *NPAS4* is an activity dependent transcription factor that has been shown to play an important role in regulating synaptic plasticity in medium spiny neurones within the nucleus accumbens^59^ and recent research has demonstrated regulation of *NPAS4* by nearby lncRNAs^60^. Taken together with the theorised role of mesolimbic dopamine circuitry in plasticity^7^, these findings suggest *NPAS4* regulation in the nucleus accumbens as a plausible candidate for future work attempting to elucidate the biological underpinnings of plasticity.

Consistent with prior research on other brain traits^35^, we found that neurones, and not glial or support cells, showed enriched SNP-heritability for the FFM personality traits across the top decile of genes expressed in the Siletti et al.^36^ superclusters. Moreover, each personality trait showed enrichment across multiple neuronal cell types which again is consistent with diffuse contributions to personality traits from across different cell types and brain regions. For stability and plasticity, none of the Siletti superclusters were significantly enriched after correction for multiple testing. This likely indicates we were underpowered for stratified Genomic SEM analyses, which are sensitive to low powered traits and small annotations.

As has been documented previously^19,20^ genetic correlation analyses indicated substantial genetic overlap between the FFM personality traits and psychiatric conditions. This was principally observed for agreeableness and inverted neuroticism which were negatively associated with psychiatric conditions, and openness, which primarily displayed moderate positive associations with the psychotic disorders (bipolar disorder and schizophrenia) and autism spectrum disorder. As suggested by the patterns at the univariate level, the meta-trait stability, which captures the shared genetic effects across agreeableness, conscientiousness and inverted neuroticism, also showed substantial genetic overlap with all psychiatric conditions tested, except for Tourette’s syndrome. Subsequent MR analyses indicated bi-directional effects between stability, neuroticism and psychopathology, with genetic propensity for low neuroticism and high stability being broadly protective of psychopathology risk and increased genetic liability for psychopathology being associated with lower stability and higher neuroticism. This is consistent with longitudinal research that suggests a bidirectional relationship between these personality traits and general psychopathology^15^ and points towards complex bi-directional pathways linking emotional stability and psychiatric risk.

It has long been noted that emotional states are directly tied to the goal related behaviour of organisms, with negative affect reflecting impeded or impaired progress towards desired states^61^. One of the key components of general psychopathology (captured by the *p*-factor) is a general tendency towards negative emotionality^28,62^. Given the meta-trait stability captures the robustness of systems supporting goal directed behaviour and emotional regulation, it is likely that genetic factors which lower stability may increase susceptibility to negative affect when goals are blocked or the environment is uncertain. Conversely, excesses of negative affect which categorise psychopathology may impair the cognitive and motivational processes required for maintaining goal directed behaviour. The observed bi-directional links between stability and psychopathology may then reflect shared pathways in how individuals maintain long term goal directed behaviour and manage affective responses to goal disruption.

There are several limitations that need to be considered when interpreting the results of the present study. Firstly, due to data availability, the present study only included data from European ancestry, and so findings may not generalise to other ancestry groups. Different measures of the FFM personality traits where also used between cohorts which may introduce measurement heterogeneity across instruments. Secondly, as the meta-trait plasticity is defined using only two indicators (openness and extraversion), constraints were applied to identify the factor. Although fit was comparable to other Genomic SEM (for example the p-factor model in the original genomic SEM paper^25^) our model fit was below the ideal “good fit thresholds” (CFI > 0.90, SRMR < 0.05). Despite boosting GWAS power for the FFM personality traits, our study was still likely underpowered for stratified Genomic SEM analyses, despite enrichment at the univariate level. This was also likely for CAUSE analyses where there is only moderate genetic overlap between traits. Furthermore, although CAUSE accounts for shared pleiotropy, causal effect estimates may still be biased by horizontal pleiotropy and residual pleiotropy may confound bi-directional effect estimates. Finally, we did not conduct any fine-mapping or colocalization analyses for GWAS or TWAS signals, nor replicate loci in an external cohort. Hence the present work should be interpreted as a broad mapping of the genetic landscape of the FFM meta-traits which highlights *potential* pathways for future follow up work. In parallel to the present work, a preprint from the Revived Genomics of Personality Consortium reporting GWAS for each of the FFM traits, including up to 1.2million participants across multiple ancestries has become available^63^. Future work should seek to replicate our findings in this larger cohort once data becomes available, given the multiple ancestries and substantially increased power it offers.

Here we leverage recent advances in GWAS of the FFM personality traits and multivariate statistical methods to perform a genomic investigation into the FFM meta-traits stability and plasticity. Using a variety of in-silico methods, we highlight potential biological pathways underpinning personality traits and characterise the genetic relationship between personality traits and psychiatric conditions. These findings importantly contribute to our knowledge of the genetic architecture of personality and how it relates to psychopathology risk.

## Methods

### GWAS meta-analysis & Multivariate GWAS

We conducted a GWAS meta-analysis of the FFM personality traits, combining the European GWAS summary statistics from Gupta et al.^20^ (N = 220,015– 623,482) and the 23andMe Research Institute sample from Lo et al.^19^ (N = 59,225). The cohorts included in the Gupta et al meta-analysis were: UK biobank^21^ (neuroticism only; N = 372,903), Genomics of Personality Consortium^22,24,64^ (N = 17,375 for agreeableness, conscientiousness and openness; N = 63,661 for extraversion and neuroticism) and Million Veterans Program^20^ (N = 220,000 – 240,000). All contributing cohorts used in our meta-analyses obtained informed consent and relevant ethics approvals. Information about the original GWAS (genotyping, imputation, ethics etc.) can be found in the original publications.

For meta-analysis of the FFM traits, the summary statistics from Gupta et al^20^ and the 23andMe Research Institute^19^ were combined using the inverse variance weighting scheme in METAL^29^. Prior to analysis, data sets were checked for alignment and filtered to exclude SNPs with an info score < 0.6 and MAF < 0.001. For the meta-traits, the *usergwas*() function in Genomic SEM^25^ was used to conduct a multivariate GWAS on stability and plasticity.

Independent GWS loci were identified using the SNP2GENE function in FUMA^65^ using the 1000 genomes (phase 3) European reference panel. Independent significant SNPs were defined as having a GWS threshold of p < 5e^-8^ and an LD threshold of r2 < 0.6 with other local SNPs. Lead SNPs were identified using a secondary LD threshold of r2 < 0.1. GWS loci were merged if lead SNPs overlapped within ±250kb of one another. The SNP2GENE function in FUMA was also used for gene mapping. Genes were mapped for each of the GWS loci identified using both positional (within ± 10kb of lead SNPs) and cis-eQTL mapping using PsychENCODE^66^, BRAINEAC^67^ and GTEX v8^40^ (all tissues) annotations. For direct comparison between studies, this FUMA pipeline was also applied to the Gupta et al. European meta-analyses to ensure consistent locus definitions. Novel loci were defined as genetic loci which did not overlap with genome wide significant loci from Gupta et al^20^.

For the meta-traits, PLINKv1.07^68^ was used for identification of QSNP loci. Lead QSNPs were identified using QSNPp < 5e^-8^, distance cutoff = 250kb, r^2^ < 0.1 using the 1000 Genomes European reference panel. Lead QSNPs and their LD partners were removed from the meta-trait summary statistics prior to running through the FUMA pipeline to remove loci which did not appear to have their effects mediated by the meta traits.

### Genetic correlations and SNP based heritability

The multivariable version of LDSC^30^ in the Genomic SEM package^25^ was used to calculate the SNP based heritability of the FFM personality traits, and the genetic correlations between the FFM personality traits and psychiatric conditions. This also produced the S and V matrices required for Genomic SEM. Genetic correlations between the meta-traits (stability and plasticity) and psychiatric conditions were estimated within Genomic SEM by specifying the meta-trait factor structure (Figure 1 B) and estimating covariances between the latent factors and psychiatric conditions^26^. The European 1000 genomes phase 3 panel was used for LD reference & GWAS summary statistics were filtered to include only HapMap 3 SNPs prior to LDSC analysis.

### Genomic structural equation modelling

Exploratory factor analysis on the genetic covariance matrix of FFM personality was conducted using the *factanal()* function in the stats R package. Both oblique and orthogonal rotations (Promax and Varimax respectively) were tested for 1 factor and 2 factor solutions. There were minimal differences in loading patterns between the rotations used (Supplementary Table 11). Results for the Varimax rotation are reported in text. Factor loadings > 0.3 were retained for subsequent Confirmatory factor analysis.

Confirmatory factor analysis was conducted using the *usermodel()* function in Genomic SEM using the diagonally weighted least squares estimator. We used benchmarks of: CFI > 0.85 and SRMR < 0.10 as acceptable fit, CFI > 0.90 and SRMR < 0.05 as good fit, and CFI > 0.95 and SRMR = 0 as great fit^25^. Unit variance identification was used for all models tested unless otherwise specified.

### Transcriptome wide association study

FUSION^41^ was used to conduct TWAS for the FFM traits and T-SEM^27^ (a multivariate extension of FUSION) was used for the meta-traits. We used the precomputed GTEX-V8^40^ expression weight panels for the 13 available brain tissues and whole blood. These models are pre filtered to retain nominally significant genes (p < 0.05) which show significant out of sample prediction (*R*^2^ > 0.01). A strict Bonferroni correction (p < 0.05/number of successfully tested gene models) for multiple testing was used in addition to a more permissive Benjamini-Hochberg FDR correction for suggestive gene-trait associations. For the meta-traits, a Bonferroni corrected Q-trait p value < 0.05 was used to identify heterogenous gene-trait associations. Of the 65,417 gene-tissue models tested for each trait, 30,601 for agreeableness and conscientiousness, 30,596 for extraversion, 31,058 for openness, and 31,257 for inverted neuroticism ran successfully.

### Cell Type Enrichment Analyses

For the FFM traits, stratified LDSC regression^69^ was used to examine enrichment for FFM SNP-based heritability across the 31 brain cell supercluster annotations from Yao et al^35^ and the baselineLD v2.2 annotations^37^. The annotations from Yao et al^35^ correspond to the top decile of gene expression for the superclusters identified in Siletti et al^36^. We first combined the Yao et al. annotations with the baselineLD v2.2 annotations and then recomputed LD scores using the 1000 Genomes reference panel, allowing us to determine heritability enrichment in the cell types independent of the baseline annotations. S-LDSC was then run 32 times for each FFM trait (baseline only and baseline plus each of the 31 supercluster annotations).

For the meta-traits, stratified Genomic SEM^26^ (multivariate extension of S-LDSC) was used across the same annotations used in the FFM traits. All continuous annotations from baselineLD v2.2 and any annotations which produced warnings for negative residual variances were excluded from interpretation. A Benjamini-Hochberg correction for multiple testing was used for all Enrichment analyses.

### Mendelian randomisation analyses

To investigate the causal relationship between personality traits and psychiatric conditions, we used the Bayesian Mendelian randomisation (MR) method CAUSE^57^, which allows comparison of null, sharing and causal models. We conducted bidirectional MR for each personality trait-psychiatric condition pair that showed significant genetic overlap in our genetic correlation analyses to test causality in both directions. CAUSE nuisance parameters were computed using 1 million random variants for each personality trait-psychiatric condition pair. Instruments for the exposure traits were obtained using clumping in PLINK^68^ with an LD r^2^ threshold of < 0.01 and a significance threshold of p < 0.001, using the 1000 Genomes Project European reference panel. The default CAUSE priors were used for all analyses. For stability and plasticity, QSNP significant loci were removed from the GWAS summary statistics prior to running CAUSE to ensure that only variants having their effect mediated by the meta-traits contributed to causal estimation (see above in GWAS meta-analysis & Multivariate GWAS methods for details about Q-SNP clumping). We used a Benjamini-Hochberg correction for multiple testing across all cause analyses (FDR adjusted p values for all CAUSE analyses are presented in supplementary table 50). We also considered nominally significant (CAUSE p < 0.05) results as being suggestive of a causal relationship.

## Acknowledgements

We would like to thank the research participants and employees of 23andMe Research Institute for making this work possible. We also thank all the research groups whose data were used in our work for making their data/code publicly available^20,35,40,46–56^. Without this, the current work would not have been possible.

This research was supported by an Australian Government Research Training Program (RTP) Scholarship doi.org/10.82133/C42F-K220. EH is funded by National Health and Medical Research Council Leadership Investigator Award (GNT 2025349).

## Data availability

23andMe Research Institute summary statistics are made available through 23andMe to qualified researchers under an agreement with 23andMe that protects the privacy of 23andMe participants. Please visit research.23andme.com/collaborate/#publication for more information. The top 10,000 SNPs for all our GWAS meta-analyses are provided in the supplementary file.

Information on where to download all other GWAS summary statistics/ annotation files used in the present study are provided in the original publications.

## Code availability

No custom code was used in the present study.

## Notes

### Competing Interest Statement

The authors have declared no competing interest.

### Funding Statement

LV was supported by an Australian Government Research Training Program
(RTP) Scholarship. EH is funded by National Health and Medical Research Council Leadership Investigator Award (GNT 2025349).

### Author Declarations

The Human Research Ethics Committee of the University of South Australia deemed the current project exempt from ethical approval (Application ID: 206516)

