## Supplementary Note for "Genetic architecture of the personality meta-traits – stability and plasticity – and their overlap with psychopathology"

**Supplementary note 1:** Manhattan plots for the GWAS meta-analyses of the FFM personality traits

**Supplementary note 2:** Q-Q plots for the GWAS meta-analyses of the FFM personality traits

**Supplementary note 3:** Summary of psychiatric condition GWASs used in our analyses

**Supplementary note 4:** Forest plots summarising effect estimates for CAUSE analyses with psychiatric conditions as exposure and personality traits as outcomes

**Supplementary note 1:**

Manhattan plots for GWAS meta-analyses of the FFM personality traits.

Agreeableness:

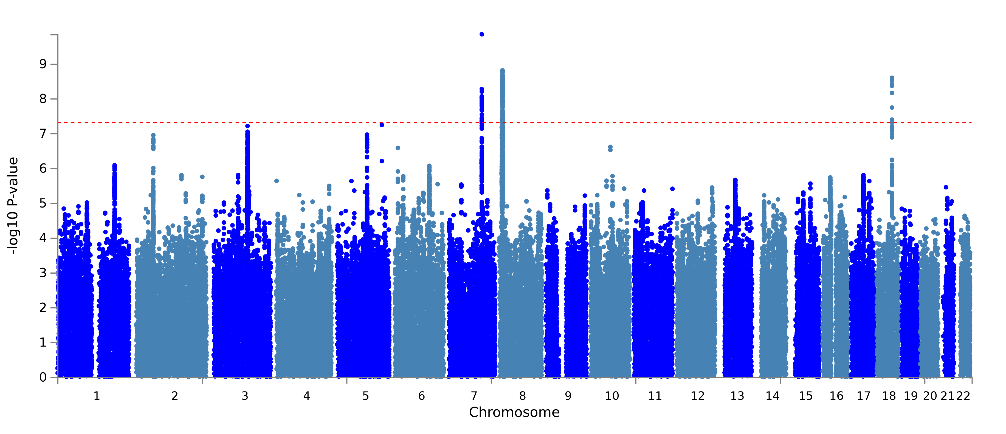

Conscientiousness:

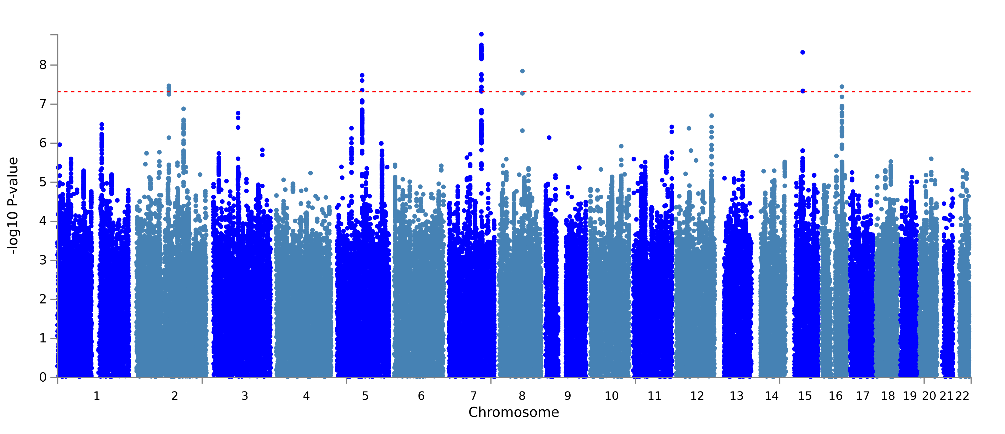

Extraversion:

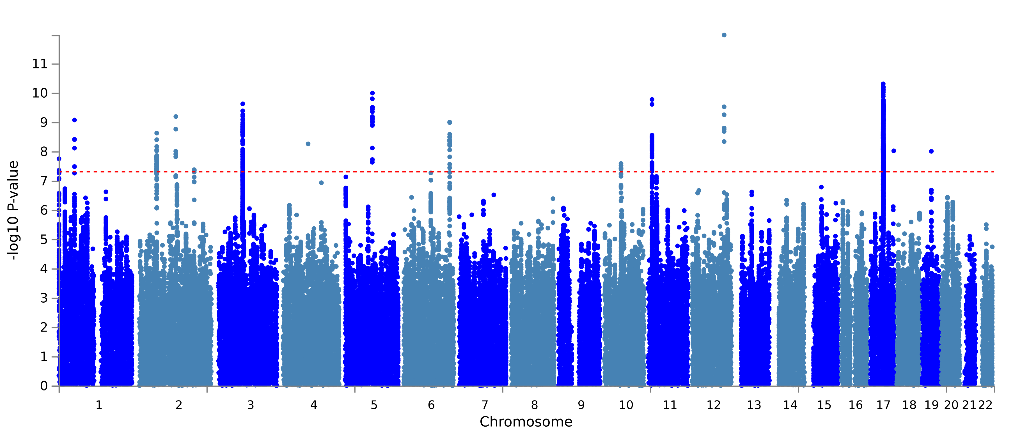

Neuroticism:

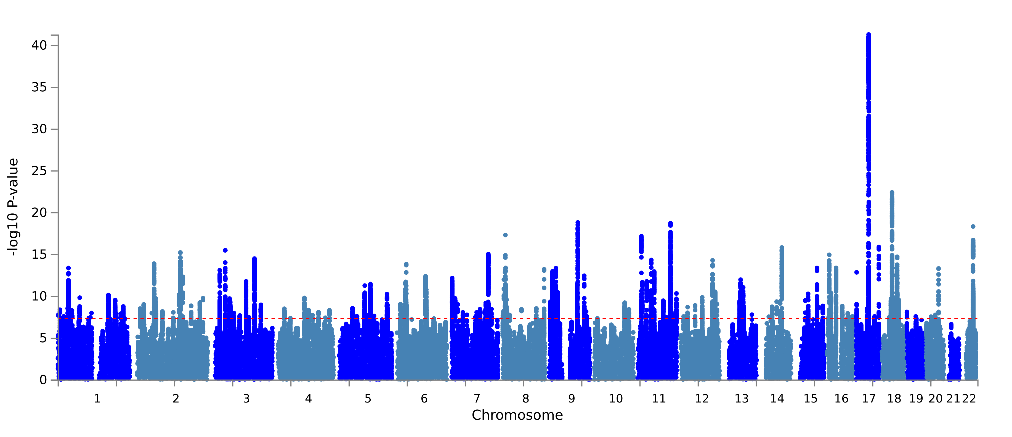

Openness:

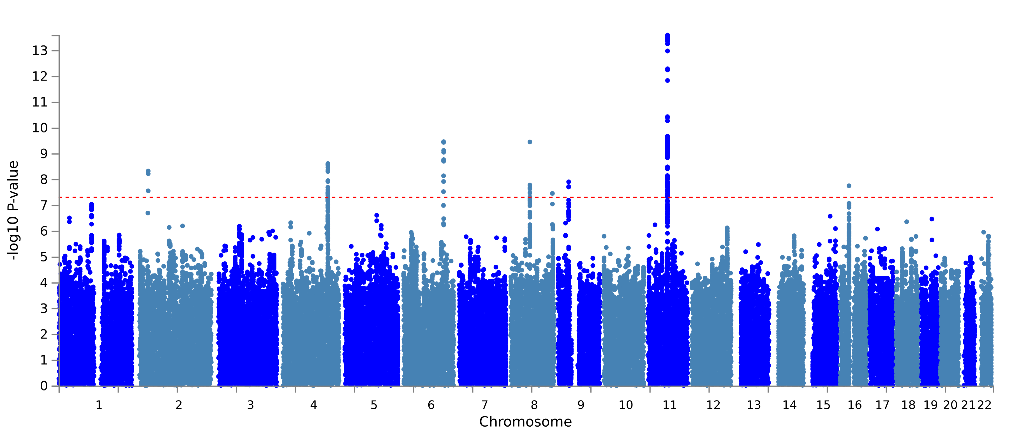

**Supplementary note 2**

Q-Q plots for GWAS meta-analyses of FFM traits and FFM meta-traits.

Agreeableness:

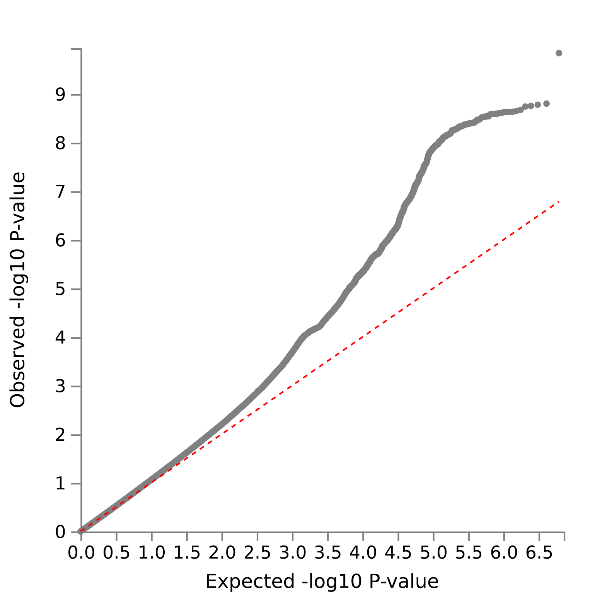

Conscientiousness:

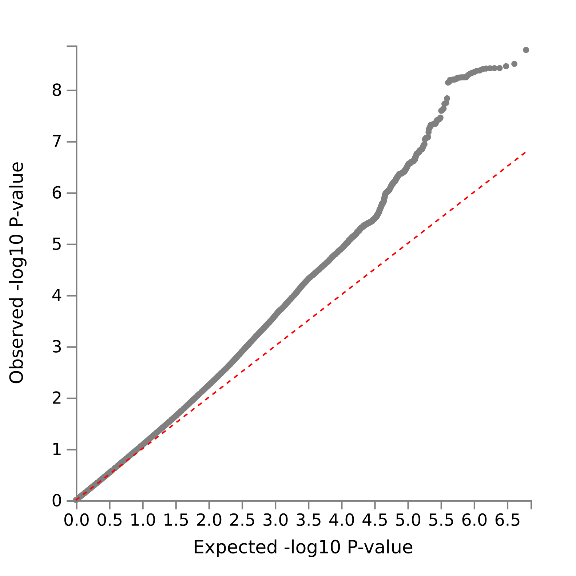

Extraversion:

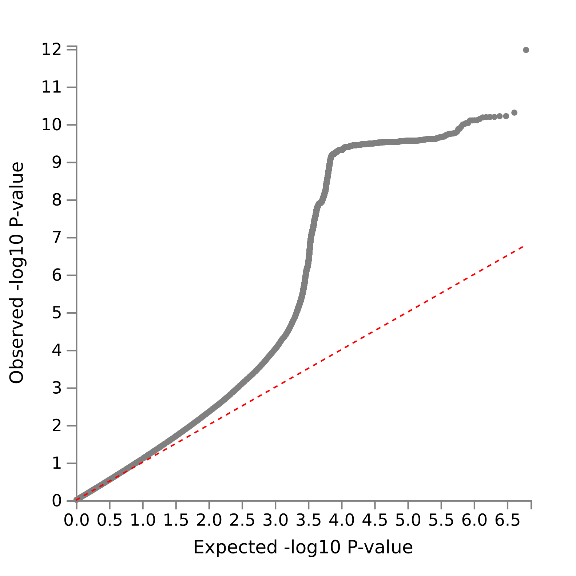

Neuroticism:

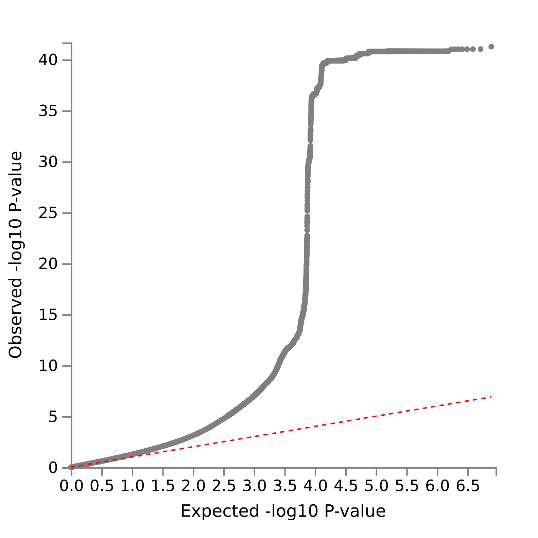

Openness:

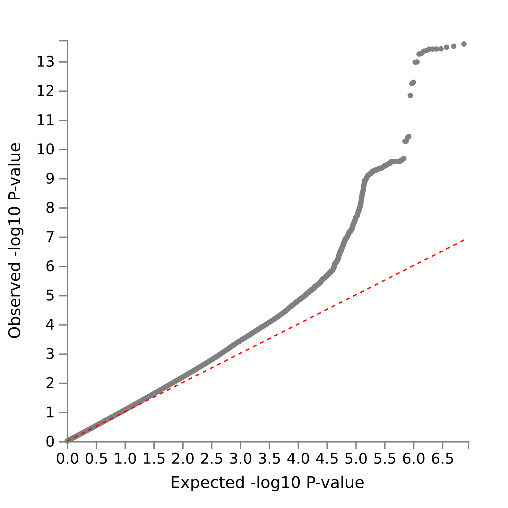

Plasticity:

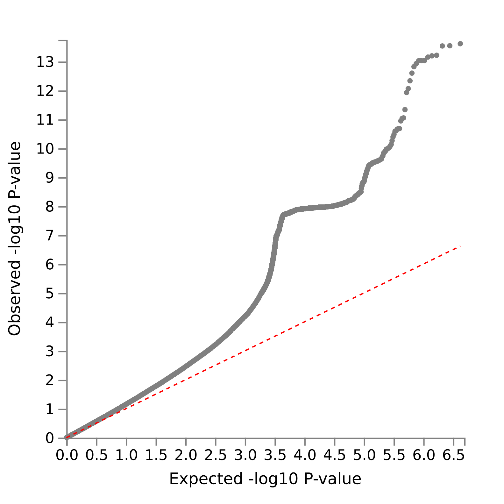

Stability:

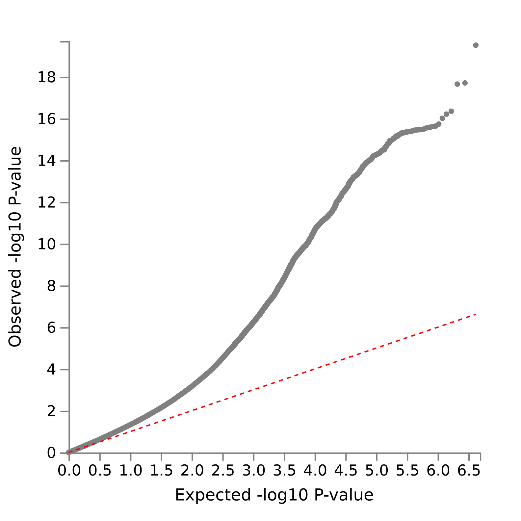

**Supplementary note 3**

Summary of the psychiatric condition genome wide associations studies used in genetic correlation analyses. For multi ancestry GWAS, only the EUR summary statistics were used.

| **Psychiatric condition** | **Population prevalence** | **Sample size (cases/controls)** | **SNP h^2^ (SE)** | **GWS loci** | **Mean Chi^2^** | **LDSC intercept** |
| --- | --- | --- | --- | --- | --- | --- |
| ADHD^1^ | 0.087 | 38691/186843 | 0.209 (0.01) | 27 | 1.449 | 1.02 |
| ALCH^2^ | 0.12 | 10206/28480 | 0.128 (0.023) | 1 | 1.058 | 1.01 |
| AN^3^ | 0.009 | 16992/55525 | 0.152 (0.01) | 8 | 1.302 | 1.02 |
| ANX^4^ | 0.31 | 1096458 | 0.053 (0.002) | 40 | 1.59 | 1.00 |
| AUT^5^ | 0.02 | 18381/27969 | 0.143 (0.012) | 5 | 1.20 | 1.01 |
| BIP^6^ | 0.01 | 131969/2322416 | 0.199 (0.008) | 229 | 1.64 | 1.06 |
| MDD^7^ | 0.21 | 170756/329443 | 0.094 (0.004) | 101 | 1.59 | 0.995 |
| OCD^8^ | 0.02 | 2688/7037 | 0.281 (0.044) | 0 | 1.05 | 0.993 |
| PTSD^9^ | 0.068 | 23212/151447 | 0.039 (0.007) | 2 | 1.08 | 1.02 |
| SCZ^10^ | 0.01 | 55193/74172 | 0.222 (0.008) | 287 | 2.05 | 1.08 |
| TS^11^ | 0.007 | 4819/9488 | 0.218 (0.252) | 1 | 1.12 | 1.01 |
| *Note. GWS loci are those reported in the original GWAS publication. SNP h^2^, mean chi^2^ and LDSC intercept are results from our LDSC analyses. We used the method recommended in Grotzinger et al.^12^ to calculate effective sample size for each GWAS.* | | | | | | |

**Supplementary note 4**

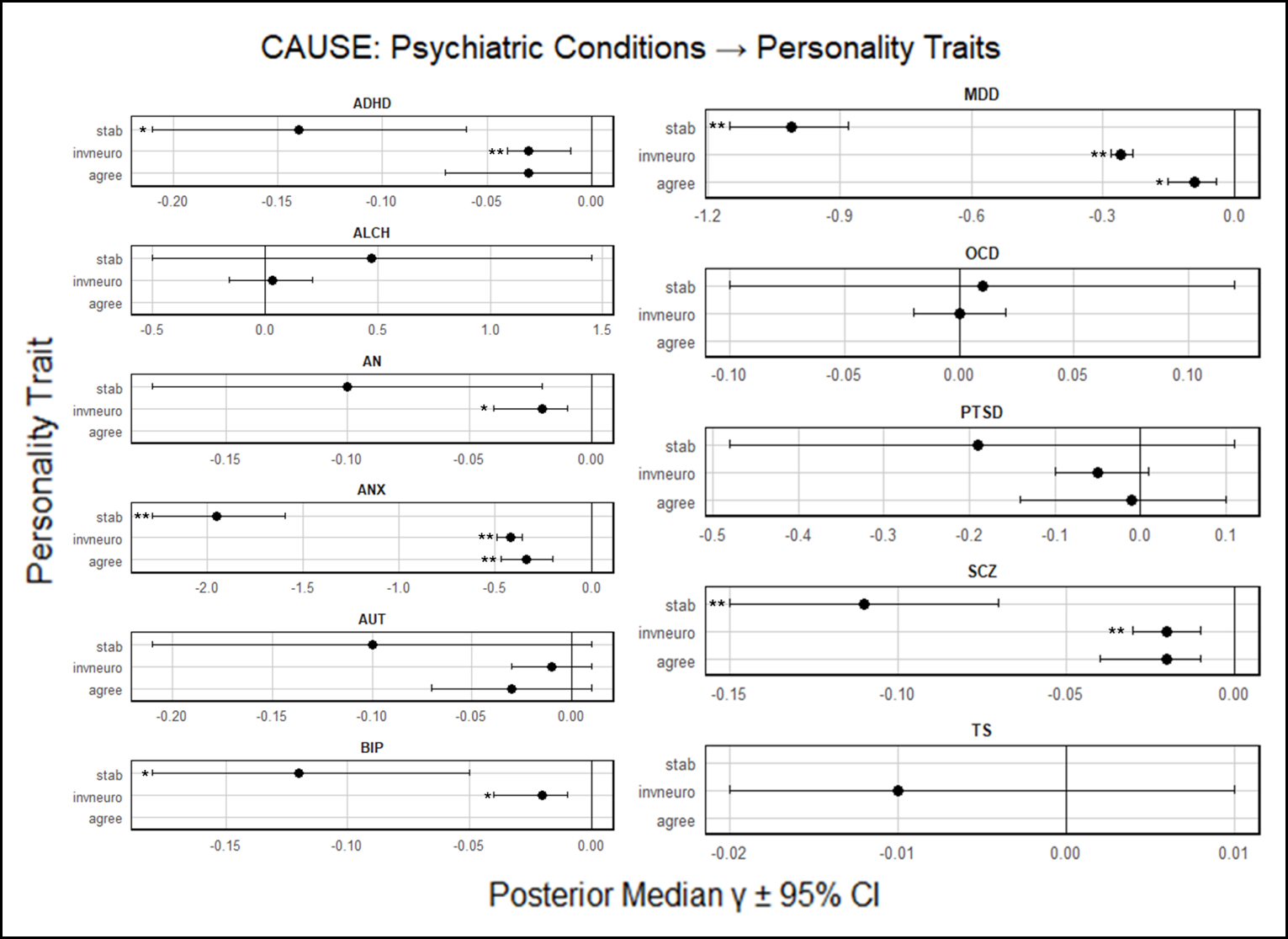
Forest plots summarising effect estimates for CAUSE analyses with psychiatric conditions as exposure and personality traits as outcomes

*Forest plots showing reverse causal effect estimates of psychiatric conditions on agreeableness, inverted neuroticism and stability conditions (posterior median γ ± 95% CI). Note: Effect sizes are scaled to the exposure trait and indicate relative causal influence, not absolute magnitude. * = nominally significant results (CAUSE p < 0.05), ** = significant results after FDR correction (FDR_BH_ < 0.05). ADHD = attention deficit hyperactivity disorder, ALCH = problematic alcohol use, AN= anorexia nervosa, ANX = anxiety, AUT = autism spectrum disorder, BIP = bi-polar disorder, MDD = major depressive disorder, OCD = obsessive compulsive disorder, PTSD = post-traumatic stress disorder, SCZ = schizophrenia, TS = Tourette syndrome, agree = agreeableness, invneuro = inverted neuroticism, stab = stability.*
